# GLP-1 Receptor Agonists and Cardiovascular Events in Adults with Obesity and Autoimmune Disease: A Target Trial Emulation

**DOI:** 10.1101/2025.11.10.25339912

**Authors:** Hao Dai, Yao An Lee, Austin Natalie, Whitney Jackson, Angela Pham, Jake Levine, Rotana Radwan, Jingchuan Guo, Jiang Bian, Amy J. Sheer

**Author notes:** The two authors equally contribute to the manuscript and serve as co-first authors (Hao Dai, Yao An Lee). **Corresponding Author:** Amy J. Sheer, MD MPH, Associate Professor of Medicine, Program Director Obesity Medicine Fellowship, Diplomat of the American Board of Obesity Medicine, Department of Medicine, University of Florida College of Medicine, Address: 1600 SW Archer Rd, Gainesville, FL 32610. **Funding source**: NIH/NIDDK R01DK133465.

## Abstract

**Importance:** Obesity and autoimmune diseases (AID) are each associated with elevated risk of cardiovascular and thromboembolic events due to chronic systemic inflammation. Glucagon-like peptide-1 receptor agonists (GLP-1RAs) have demonstrated cardiovascular and metabolic benefits in patients with type 2 diabetes and obesity, but their effects in patients with obesity and comorbid AID remain uncertain.

**Objective:** To evaluate the association between GLP-1RA use and the risk of major adverse cardiovascular and thromboembolic events among adults with obesity and AID eligible for anti-obesity medication (AOM) therapy.

**Design:** This retrospective cohort study emulated a target trial using 2014-2024 electronic health record data from the OneFlorida+ network, which includes 21 million individuals across Florida, Georgia, and Alabama. Adults with obesity and AID who met AOM eligibility criteria were included. Propensity score matching (1:1) was applied using a time-dependent framework to balance baseline covariates between GLP-1RA users and non-users.

**Participants:** AID Adults (≥18 years) who were eligible for AOM treatment.

**Exposure:** GLP-1RA use versus non-use.

**Main Outcomes and Measures:** The primary outcomes were myocardial infarction, stroke or transient ischemic attack (TIA), pulmonary embolism (PE), venous thromboembolism (VTE), and coronary revascularization. Secondary outcomes included hospitalization, emergency department (ED) visits, and all-cause mortality.

**Results:** The matched cohort included 13,204 GLP-1RA users and 13,204 non-users (mean age, 54.7 ± 14.5 years; 73.4% female; mean BMI, 37 kg/m²). Compared with non-users, GLP-1RA users had lower incidence rates (per 1000 person-years) of PE (6.4 vs 9.5), VTE (16.6 vs 20.4), and mortality (9.5 vs 16.9). GLP-1RA use was associated with lower hazard of stroke/TIA (HR, 0.87 [95% CI, 0.76-0.99]; P = .039), PE (HR, 0.69 [95% CI, 0.56-0.86]; P = .001), VTE (HR, 0.83 [95% CI, 0.72-0.95]; P = .007), ED visits (HR, 0.79 [95% CI, 0.75-0.83]; P = .000), and mortality (HR, 0.56 [95% CI, 0.47-0.66]; P = .000).

**Conclusions and Relevance:** Among adults with obesity and AID, GLP-1RA use was associated with reduced thromboembolic events, lower emergency department utilization, and decreased mortality. These findings suggest potential cardiovascular and survival benefits of GLP-1RAs in a high-risk, understudied population.

**Key Points:** *Question:* Is the use of glucagon-like peptide-1 receptor agonists (GLP-1RAs) associated with cardiovascular and thromboembolic outcomes among adults with obesity and autoimmune disease (AID)?

*Findings:* In this target trial emulation using 2014-2024 electronic health record data from the OneFlorida+ network, 13,204 GLP-1RA users were compared with 13,204 matched non-users. GLP-1RA use was associated with significantly lower risks of pulmonary embolism, venous thromboembolism, emergency department visits, and all-cause mortality, with marginal associations for stroke or transient ischemic attack.

*Meaning:* Among adults with obesity and AID, GLP-1RA therapy may confer thromboembolic and survival benefits without increasing cardiovascular risk.

## INTRODUCTION

Obesity is a growing public health crisis worldwide. Per the CDC, the prevalence of obesity in the U.S. increased from 30.5% in 1999-2000 to 41.9% in 2017-March 2020,^1^ with future trends anticipating a continued surge. Patients with obesity are widely recognized to be at increased risk for cardiovascular and thromboembolic events including myocardial infarction, cerebrovascular accident, and venous thromboembolism. This elevated risk is partly mediated by chronic low-grade inflammation as excess fat accumulation (adiposity) encourages a pro-inflammatory environment, leading to endothelial dysfunction and a prothrombotic state.^2^ Given this association, risk of these complications is expected to be compounded in patients with comorbid autoimmune disease (AID).

Autoimmune diseases are estimated to affect about 15 million individuals in the United States alone, about 4.6% of the US population.^3^ These conditions are associated with persistent inflammatory responses due to frequent stimulation of the body’s immune response.^4^ Multiple studies have demonstrated that those with AID are at 40-100% higher risk of cardiovascular and thromboembolic complications compared to match controls because of this described inflammatory state.^5^

A class of medications known as glucagon-like peptide-1 receptor agonists (GLP-1RAs) have emerged as highly effective treatment options for weight reduction, glycemic control, and cardiometabolic risk reduction. Randomized controlled trials (RCTs) have shown that GLP-1RAs reduce the incidence of major adverse cardiovascular events (MACE) in patients with type 2 diabetes.^6^ More recently, the SELECT trial demonstrated similar benefit in patients who are overweight or obese with preexisting cardiovascular disease.^7^ While it is suspected that the evidence-based benefit outlined above could provide benefit for other patient populations, there is currently insufficient data to support these speculations, especially in patients with obesity and comorbid AID. In fact, patients with AID are commonly excluded from clinical trials due to their elevated risk of cardiovascular and thromboembolic complications^5^ despite being among those who would likely benefit most from interventions targeting these issues.

Emerging evidence suggests that GLP-1RAs have anti-inflammatory properties and may modulate the immune system.^9^ In the STEP 1, 2, and 3 trials, GLP-1RA therapy resulted in an overall reduction in baseline C-reactive protein (CRP) in patients with overweight/obesity using GLP-1RA therapy compared to non-users, further suggesting an anti-inflammatory benefit for this class of medications.^9^

Clinical data is necessary to bridge the evidence gap between GLP-1RA therapy and benefits in broader populations. At this time, there is little focused research evaluating how GLP-1RA therapy could lower rates of cardiovascular and thromboembolic events in patients with obesity and comorbid autoimmune diseases. Given the lack of data and potential untapped use of these medications in improving morbidity and mortality in this group, we conducted a target trial emulation study using a large real-world dataset (OneFlorida+) to investigate the association between GLP-1RA initiation and the risk for major adverse cardiovascular and thromboembolic events in patients with obesity and AID.

## METHODS

### Study Design and Data Source

We conducted a target trial emulation using a retrospective cohort study with a new-user design, applied a time-dependent propensity score matching approach to balance baseline covariates between comparison groups, thereby mimicking randomization. An intention-to-treat analysis was conducted to investigate the association between GLP-1RAs and major adverse cardiovascular and thromboembolic events among adults with obesity and AID. This study was approved and exempted of consent by the University of Florida Institutional Review Board.

Our study utilized real-world OneFlorida+ data, covering the period from 2014 to 2024. OneFlorida+ consists of 14 health care organizations and contains longitudinal patient-level EHRs for 21 million individuals from Florida, Georgia, and Alabama.

### Study Population

The study included patients eligible for anti-obesity medications (AOMs) between January 1, 2014, and January 31, 2024. According to 2013 AHA/ACC/TOS guideline, which is applied to now, AOM eligibility was defined as (1) having a recorded diagnosis of obesity (BMI ≥ 30 kg/m²) or (2) a BMI of 27-29.9 kg/m² with at least one weight-related comorbidity^10^. Patients were excluded if they were younger than 18 years old, had an AID diagnosis on the cohort entry date.

The exposure of interest was the initiation of GLP-1RA treatment (e.g., liraglutide, semaglutide, and tirzepatide, identified using RxNorm identifiers, **eTable 1 in Supplement**). The comparator group, defined as patients who did not initiate GLP-1RA therapy during the study period, was constructed by aligning non-users to GLP-1RA users within ±1 week window of the initiation date, to ensure temporal comparability of treatment assignment. The index date was the first GLP-1RA prescription date for users and the matched visit date for non-users. Patients were excluded if they had no encounter in the year before index, had <30 days of follow-up, or had prior AID diagnosis.

### Study Outcomes and Follow-up

The primary outcome was myocardial infarction, stroke/ transient ischemic attack (TIA), pulmonary embolism (PE), venous thromboembolism (VTE), and coronary revascularization. The secondary outcomes included any Hospitalization, any ED visit, and mortality. All the clinical outcomes were identified using ICD-9 and ICD-10 diagnosis codes (**eTable 2 in Supplement**).

Patients’ follow-up began at the index date and continued until the earliest occurrence of one of the following censoring events: the outcome of interest, death, loss to follow-up (the date of the last recorded clinical encounter), or the end of the study period (January 31, 2024).

### Covariates

Baseline covariates included demographics, comorbidities, medications, and laboratory values, detailed in **Table 1**. Race/ethnicity categories included Hispanic, non-Hispanic White, non-Hispanic Black, and Other (e.g., multiracial, Asian, Native Hawaiian or Other Pacific Islander, and American Indian or Alaska Native). Medications information was collected during the year before or on the index date and comorbidities were collected one year prior to the index date.

**Table 1.**
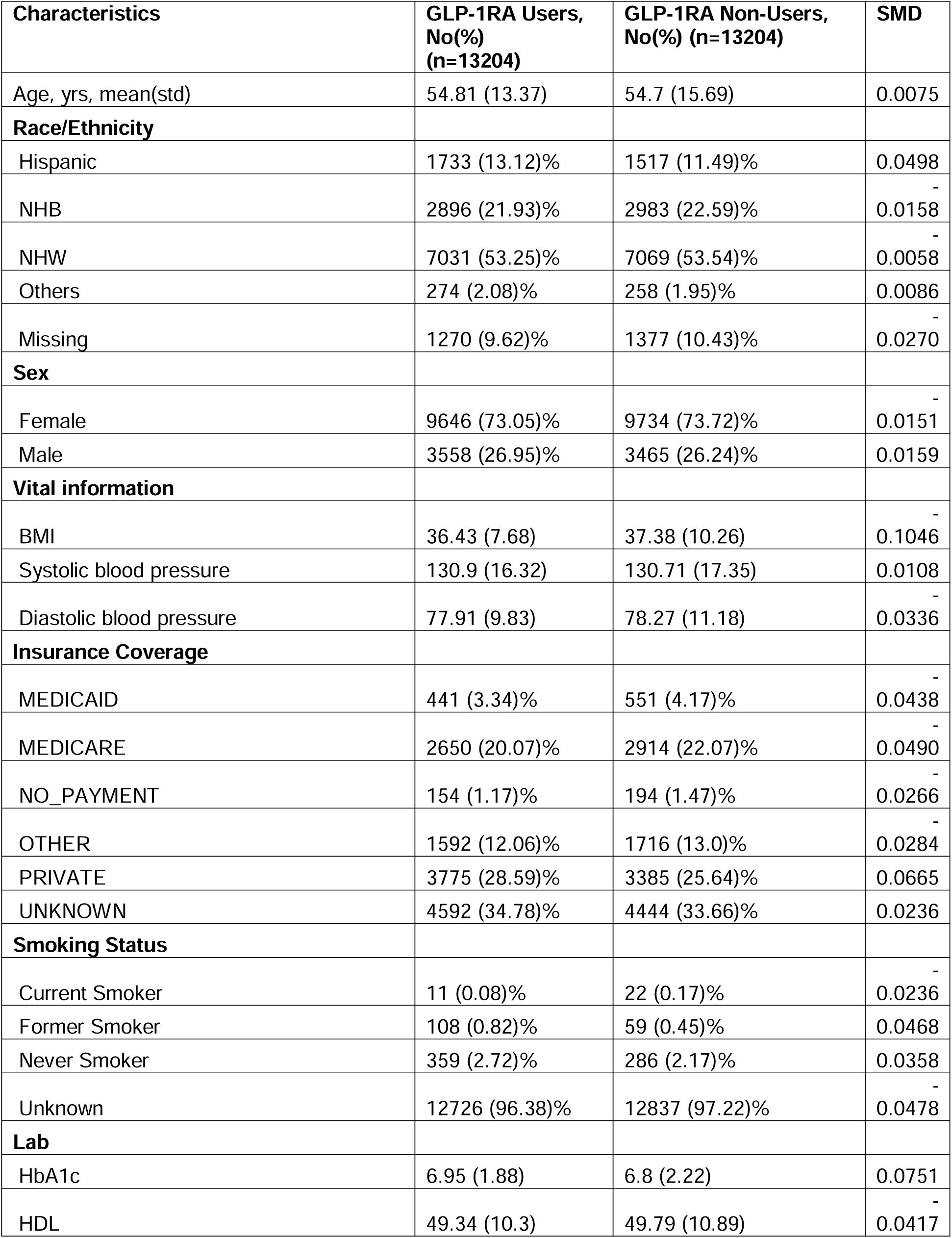
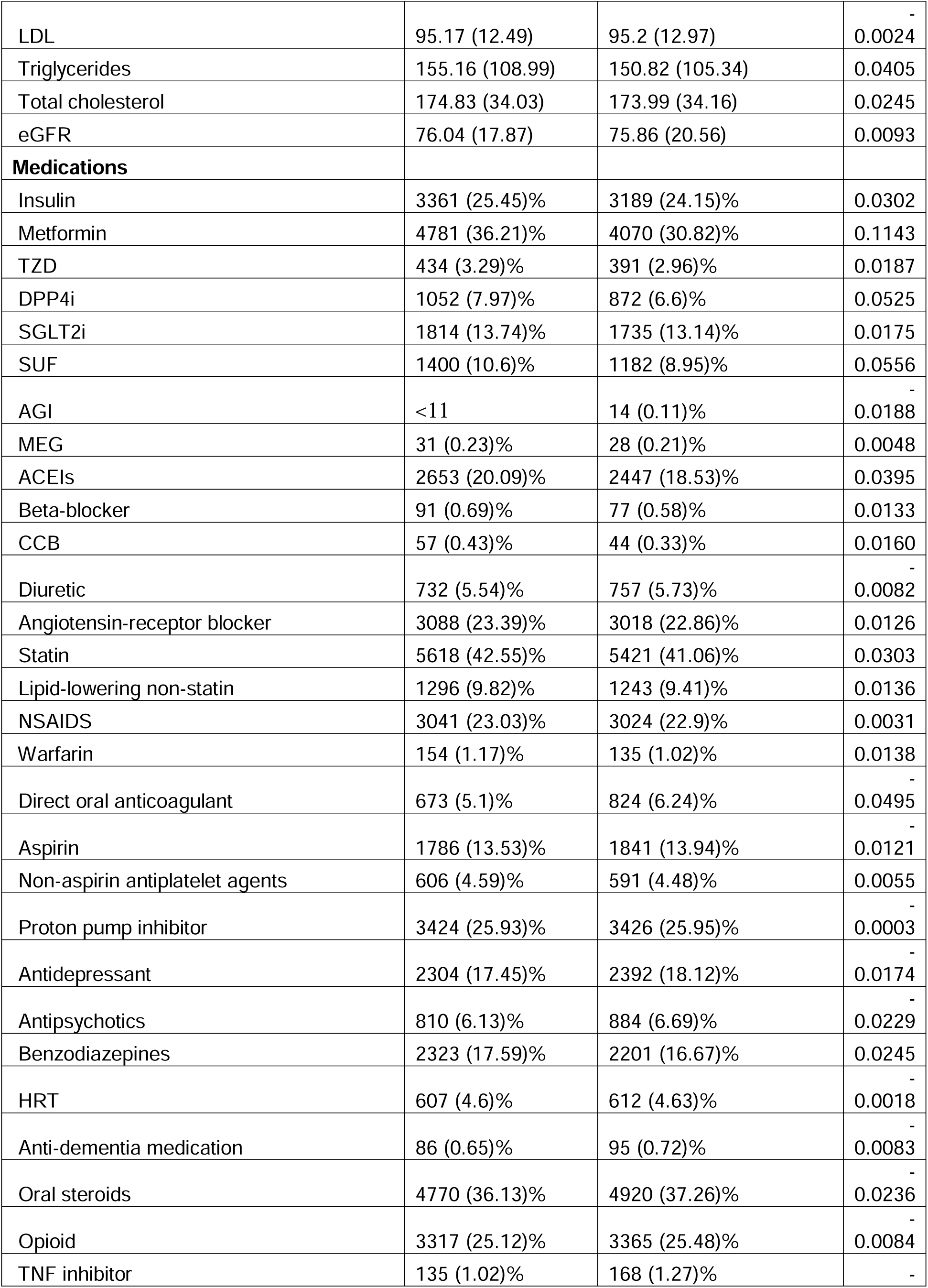
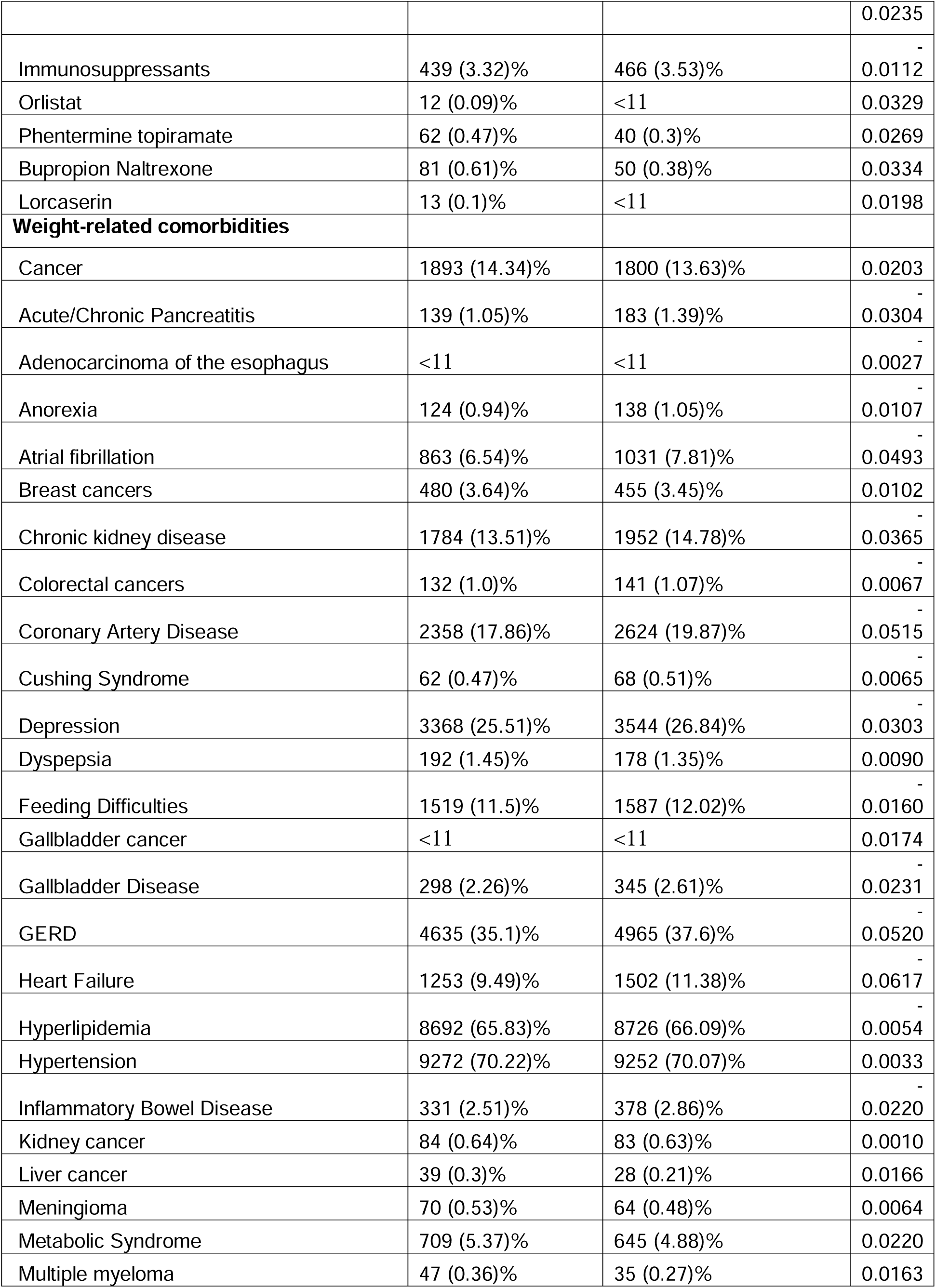
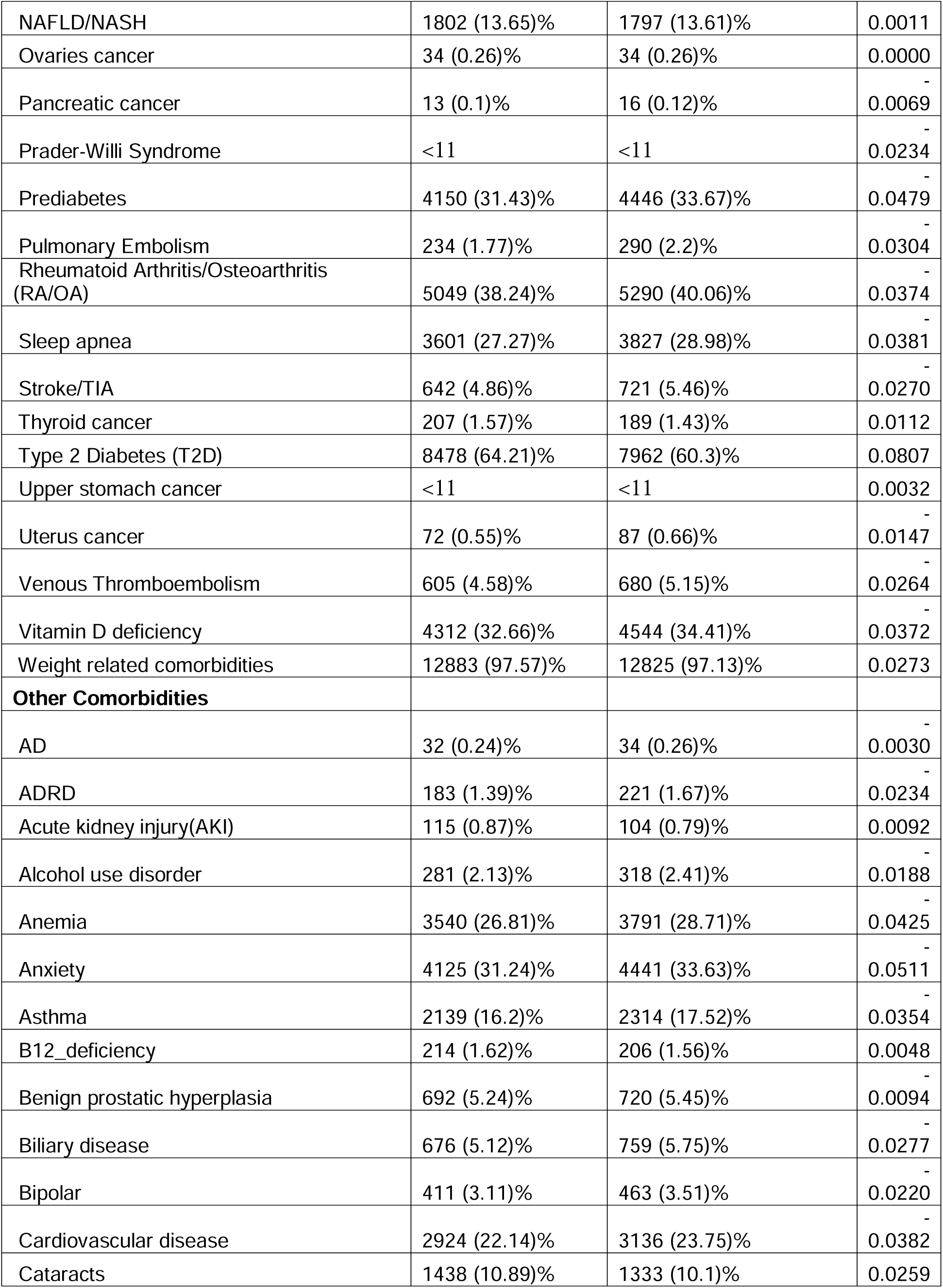
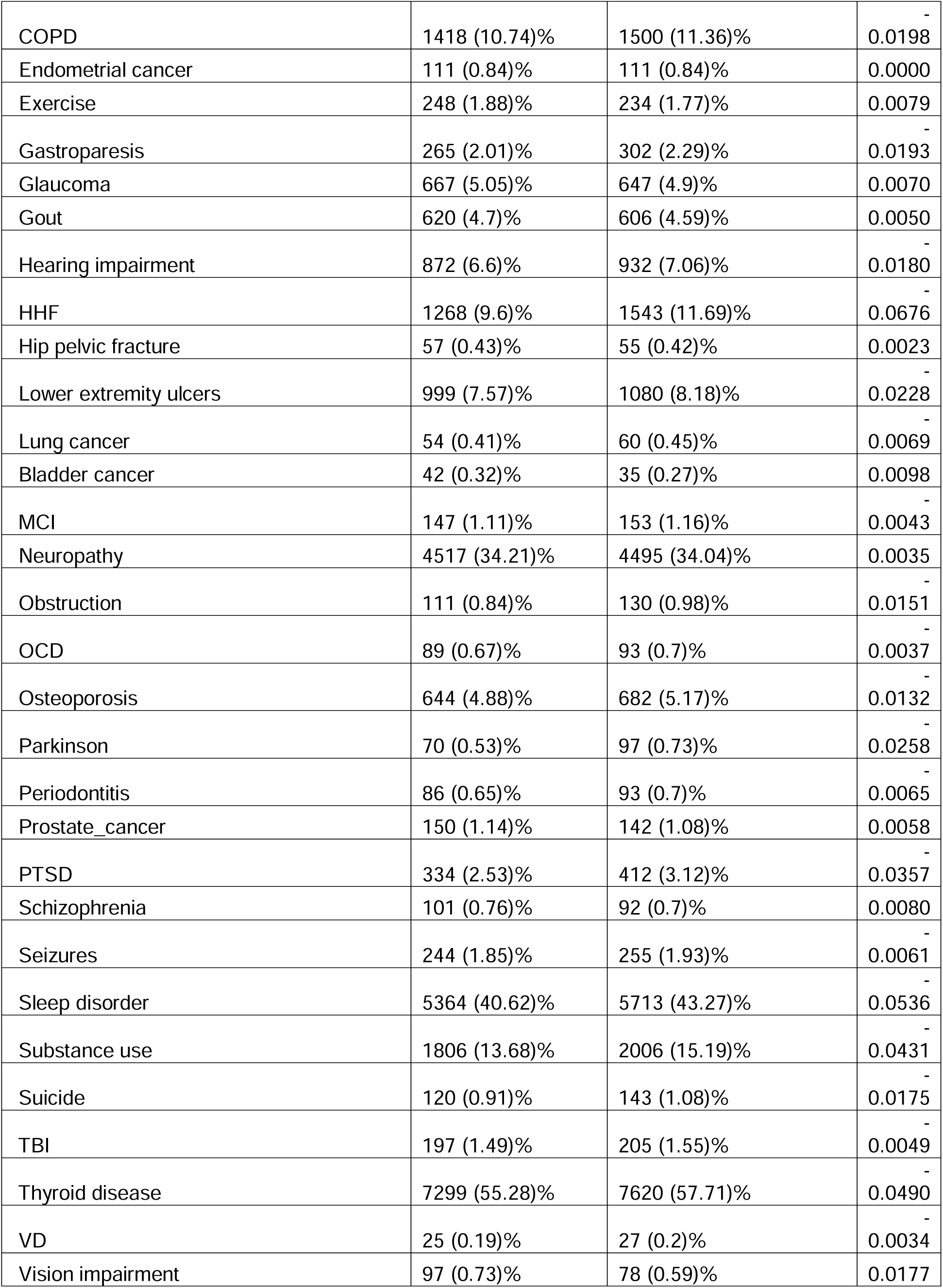
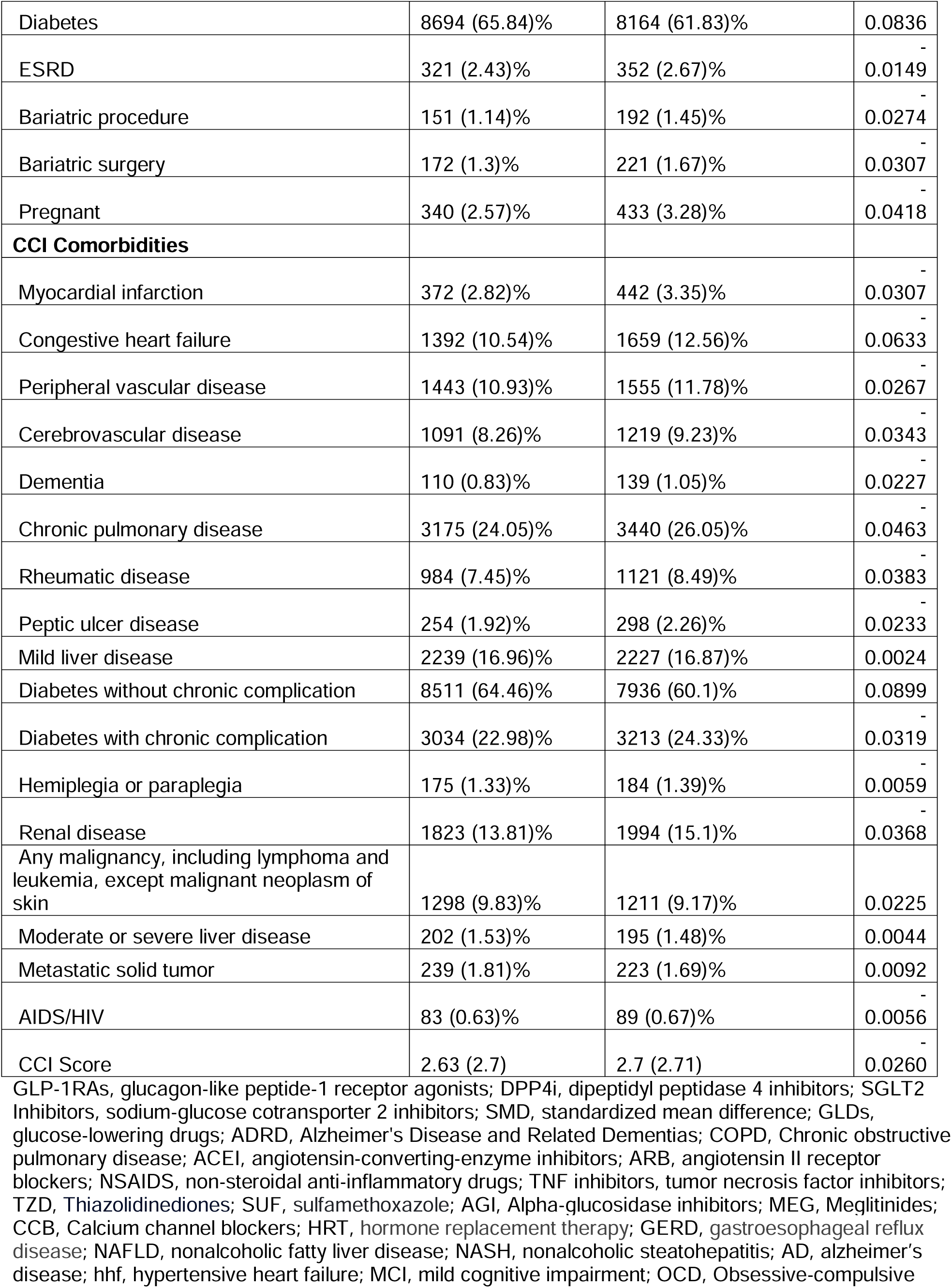
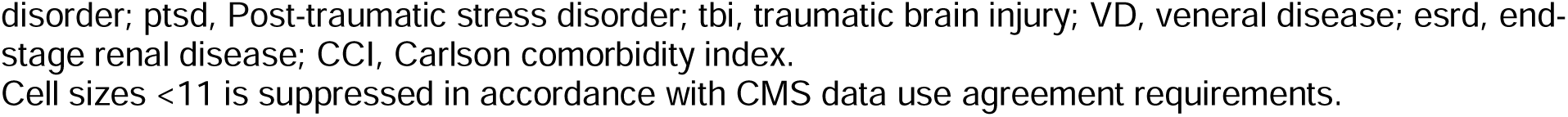
Baseline characteristics of GLP-1RA users vs. non-users after 1:1 propensity score matching.

### Statistical Analysis

To address potential confounding and enhance comparability between two treatment groups, we implemented a time-dependent 1:1 propensity score (PS) matching strategy.^11^ Propensity scores were derived from a multivariable logistic regression model incorporating a comprehensive set of baseline covariates. Matching was performed using a nearest-neighbor (KNN) algorithm without replacement and a caliper of 0.2, ensuring that each GLP-1RA user was matched to a non-user with the closest propensity score within the specified threshold.^12^ Covariate balance between matched pairs was evaluated using standardized mean differences (SMDs), with values below 0.1 considered acceptable balance between GLP-1RA users and non-users.

We calculated the incidence rate (IR) of study outcomes and employed Kaplan-Meier survival analysis to illustrate the cumulative incidence per 1000 person-years over time between the matched groups. Additionally, Cox proportional hazard regression models were used to estimate hazard ratios (HRs) with 95% confidence intervals (CIs) by comparing GLP-1RAs users vs. non-users.

### Subgroup Analysis

Subgroup analyses were conducted for prespecified factors. The prespecified subgroups included: age groups (<40, 40-64, and ≥65 years), sex (male vs. female), racial and ethnic categories (non-Hispanic White, non-Hispanic Black, Hispanic, and other groups), obesity severity (overweight vs. obesity), baseline AID by organ system (disease of endocrine system, inflammatory arthritis, vasculitis, connective tissue disorders, disease of skin system, hematological diseases, disease of nervous system, disease of digestive system and others), and individual GLP1-RA drugs (liraglutide, semaglutide, and tirzepatide).

## RESULTS

The flowchart of patient selection is presented in **Figure 1**. The study population comprised 484,467 AID patients eligible for AOM treatment in OneFlorida+ from 2014-2024, including 18,044 GLP-1RA users and 466,423 GLP-1RA non-users. After applying 1:1 PS matching (**eFigure 1 & 2 in Supplement**), the final cohort included 26,408 matched participants (13,204 GLP-1RA users vs. 13, 204 non-user). After PS matching, baseline characteristics showed a balanced distribution of key covariates. In the matched cohort (**Table 1**), mean age was 54.7 (± 14.5) years, 73.4% were female, and 53.4% were non-Hispanic White. The mean BMI at baseline was 37 kg/m^2^. The two treatment groups showed similar distributions in baseline comorbidities, medications, and other clinical factors.

**Figure 1.**
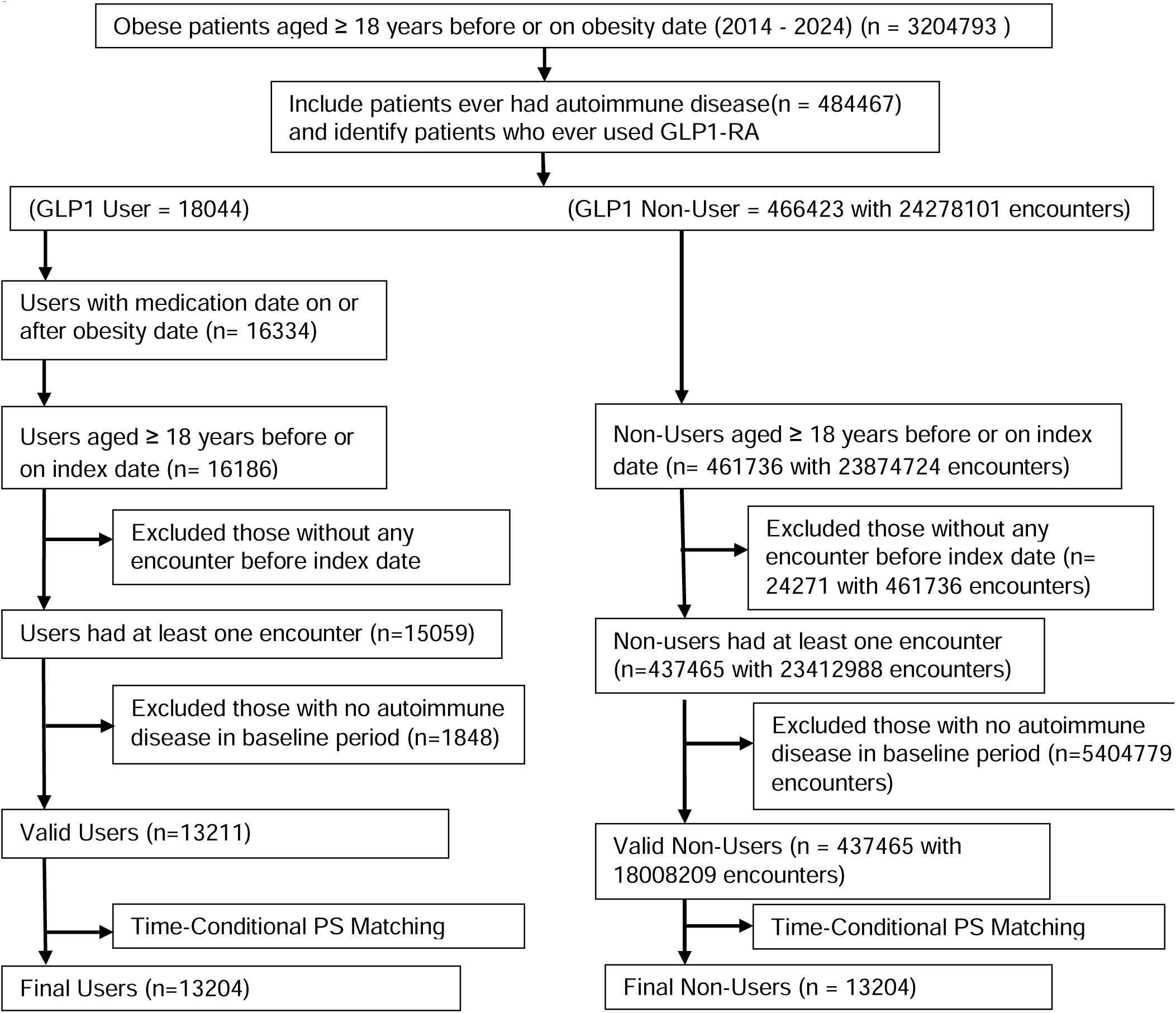
Study Group Selection Flow Diagram.

**eFigure 3** presents Kaplan-Meier plots showing the cumulative incidence of outcomes during the follow-up. Among the primary outcomes, the IR of MI was 11.2 per 1,000 person-years in GLP-1RA users compared with 13.1 in non-users, and stroke/TIA occurred at an IR of 18.8 versus 21.9 per 1,000 person-years. Pulmonary embolism showed a lower incidence among GLP-1RA users (6.4 vs. 9.5 per 1,000 person-years), as did venous thromboembolism (16.6 vs. 20.4 per 1,000 person-years). The IR of coronary revascularization was similar between the two groups (4.5 vs. 4.3 per 1,000 person-years).

For the secondary outcomes, hospitalization occurred at an IR of 210.4 per 1,000 person-years among GLP-1RA users and 231.3 among non-users, while ED visits were less frequent in the GLP-1RA group (150.9 vs. 193.0 per 1,000 person-years). Mortality showed the most notable difference, with an IR of 9.5 in GLP-1RA users compared with 16.9 per 1,000 person-years in non-users.

The HR forest plot for outcomes is shown in **Figure 2**. Specifically, GLP-1RA use was associated with a lower hazard of pulmonary embolism (HR, 0.69 [95% CI 0.56-0.86]; P = .001), venous thromboembolism (HR, 0.83 [95% CI, 0.72-0.95]; P = .007), and stroke/TIA (HR, 0.87 [95% CI 0.76-0.99]; P=.039) significantly. No significant difference was observed for myocardial infarction (HR, 0.86 [95% CI 0.72-1.02]; P = .081) and coronary revascularization (HR, 1.03 [95% CI 0.78-1.37]: P=.820). For the secondary outcomes, GLP-1RA use was linked to a lower hazard of ED visits (HR, 0.79 [95% CI, 0.75-0.83]; P = .000) and mortality (HR, 0.56 [95% CI, 0.47-0.66]; P = .000), while the hazard of hospitalization was comparable between groups (HR, 0.96 [95% CI 0.92-1.00]: P=.067).

**Figure 2.**
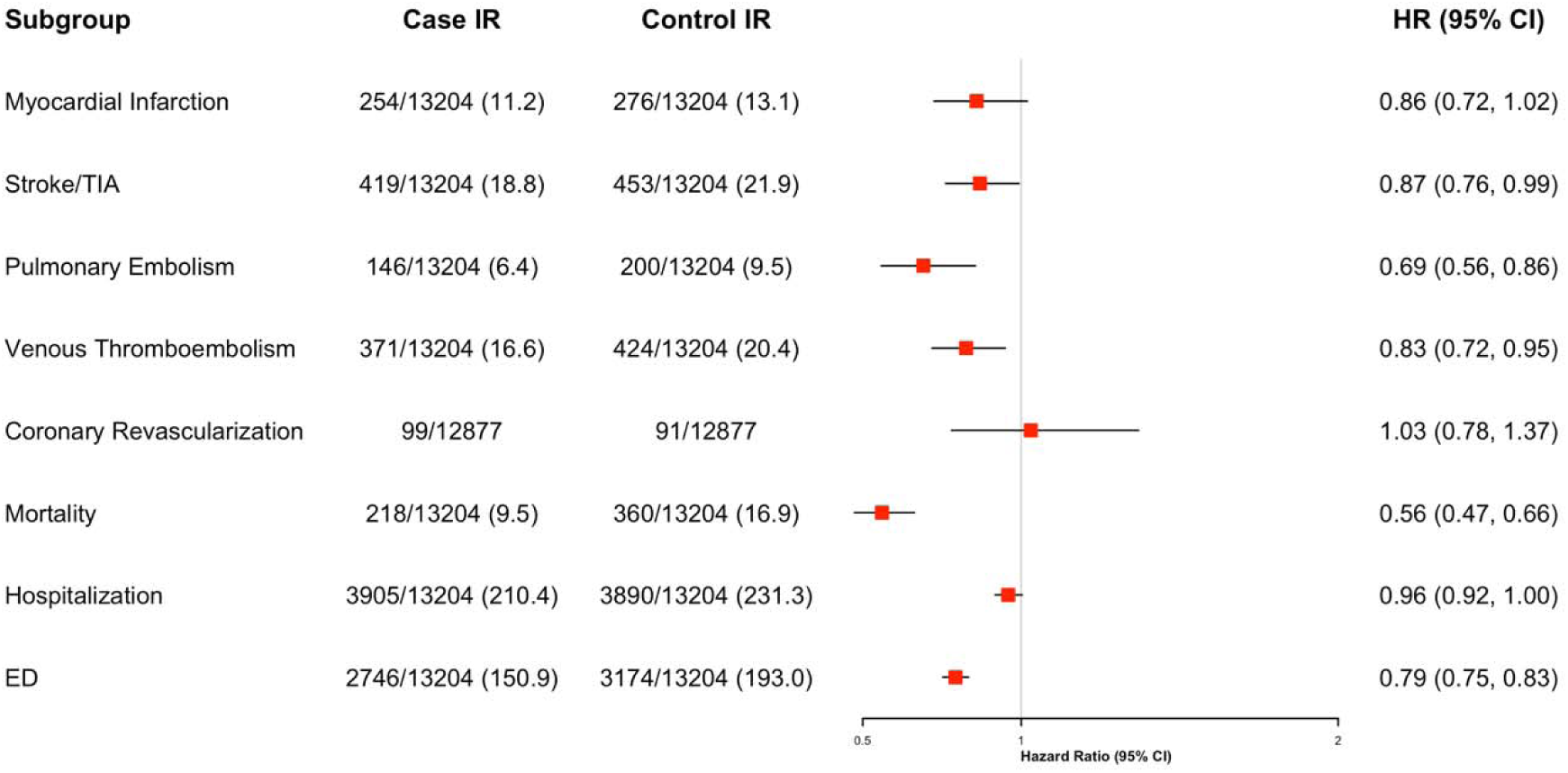
Risk of Outcomes in Patients Receiving GLP-1RAs Compared With Patients Not Receiving GLP-1RAs.

Results of subgroup analysis can be seen in **eFigure 4**. Most subgroups demonstrate a protective tendency and a reduced risk for MI, Stroke/TIA, PE, VTE, ED visits, and mortality.

## DISCUSSION

Our study found that use of GLP-1RA therapy in patients with AID was associated with statistically significant reductions in thromboembolic events (VTE and PE), emergency department utilization, and, most strikingly, all-cause mortality when compared with matched non-users. These benefits were observed across subgroups, further strengthening our confidence in these findings. While previous investigations have proven the utility of GLP-1RA therapy in patients with obesity and type 2 diabetes,^6^ ours is among the first to examine their value and potentially expand their use to those with comorbid AID.

First, our analysis revealed that GLP-1RA therapy resulted in a 31% reduction in VTE and 17% reduction in PE. This protective effect is likely multifactorial and related to the immunomodulatory and antithrombotic actions of GLP-1RAs.^13^ While prior analyses investigating the role of GLP-1RA therapy in thromboembolic risk reduction in other patient populations have delivered mixed results, our study demonstrated a clear benefit, further underscoring the utility of these medications in this high-risk group.^14^ Despite similar proposed mechanisms of risk reduction, our study demonstrated only a modest decrease in stroke/TIA risk, a nonsignificant trend for myocardial infarction, and no significant difference in coronary revascularization. This contrasts with multiple meta-analyses and RCTs that have illustrated a robust association between GLP-1RA use and MACE reduction particularly in those with type 2 diabetes mellitus and established cardiovascular disease.^15^ The reason for this variability in protective benefit is unclear but could be related to differences in pathophysiology and baseline risk.^16^

Another noteworthy discovery was the mitigation of healthcare utilization, particularly a 21% lower risk of emergency department encounters. Similar outcomes have been displayed in former studies in patients with diabetes mellitus and chronic kidney disease, which is believed to be secondary to improved cardiometabolic parameters resulting in lower rates of acute complications.^17^ This is particularly important given the substantial growth in ED-related spending accounting for roughly 5% of annual US healthcare costs equating to approximately $80 billion.^18^ This reduction in utilization did not translate to hospitalizations with similar rates of admissions among both GLP-1RA users and nonusers. The source of this discrepancy is not fully elucidated but could be explained by the often-multifactorial nature of acute decompensation in patients with more advanced disease which GLP-1RA use is unable to completely address.^19^ Nonetheless, these findings suggest that GLP-1RA use could reduce disease burden for both patients and the healthcare system.

Finally, and arguably the most clinically significant finding, was a 44% mortality reduction among patients with obesity and comorbid AID prescribed GLP-1RA therapy. This observation is further supported by previous research demonstrating reductions in all-cause mortality ranging from 37-52% in patients with diabetes^20,21^; however, our research provides evidence that this mortality benefit may extend to additional high-risk patient populations.

The results of our study should be interpreted with several limitations in mind. First, as an observational study, our analysis is susceptible to confounding, despite rigorous propensity score matching. Unmeasured factors, such as socioeconomic status, disease activity, and use of immunosuppression medications could have influenced outcomes. Second, the study looked at composite outcomes from a large data set, OneFlorida+, so specifics such as reason for all-cause hospitalization, were not ascertained. In addition, there were some patients who switched between multiple GLP-RA1 based therapies, and this effect was not accounted for. Lastly, the degree of weight change between these groups was not assessed.

## CONCLUSION

In summary, GLP-1RA use among patients with AID was associated with lower risks of VTE, PE, ED visits, and all-cause mortality, with suggestive benefits for MI and stroke. This data suggests important areas of benefit for GLP-1RA therapy in an understudied, high-risk population, who are at heightened risk due to systemic inflammation. Future studies are needed to investigate the mechanisms underlying these associations and evaluate the role of GLP-1RA as a treatment and preventative therapy in systemic inflammatory conditions.

## Data Availability

Data set Available through OneFlorida+ Clinical Research Network (email)

## Author Contributions

J. Guo and Y. Lee had full access to all the data in the study and take responsibility for the integrity of the data and the accuracy of the data analysis.

Concept and design: A. Sheer, J. Guo, and J. Bian.

Acquisition, analysis, or interpretation of data: All authors.

Drafting of the manuscript: H. Dai, Y. Lee, A. Natalie, and W. Jackson.

Critical review of the manuscript for important intellectual content: All authors.

Statistical analysis: H. Dai and Y. Lee

Obtained funding: J. Guo.

Supervision: A. Sheer, J. Guo and J. Bian.

## Reference

1. Centers for Disease Control and Prevention. Adult obesity facts. CDC. May 14, 2024. Accessed September 14, 2025. https://www.cdc.gov/obesity/adult-obesity-facts/index.html

2. Powell-Wiley TM, Poirier P, Burke LE, et al. Obesity and cardiovascular disease: A scientific statement from the American heart association. Circulation. 2021;143(21):e984–e1010.

3. Abend AH, He I, Bahroos N, et al. Estimation of prevalence of autoimmune diseases in the United States using electronic health record data. J Clin Invest. 2024;135(4). doi:10.1172/JCI178722

4. Xiang Y, Zhang M, Jiang D, Su Q, Shi J. The role of inflammation in autoimmune disease: a therapeutic target. Front Immunol. 2023;14:1267091.

5. Conrad N, Verbeke G, Molenberghs G, et al. Autoimmune diseases and cardiovascular risk: a population-based study on 19 autoimmune diseases and 12 cardiovascular diseases in 22 million individuals in the UK. Lancet. 2022;400(10354):733–743.

6. American Diabetes Association Professional Practice Committee. 10. Cardiovascular disease and risk management: Standards of care in diabetes-2025. Diabetes Care. 2025;48(1 Suppl 1):S207-S238.

7. Lincoff AM, Brown-Frandsen K, Colhoun HM, et al. Semaglutide and cardiovascular outcomes in obesity without diabetes. N Engl J Med. 2023;389(24):2221–2232.

8. Bendotti G, Montefusco L, Lunati ME, et al. The anti-inflammatory and immunological properties of GLP-1 Receptor Agonists. Pharmacol Res. 2022;182(106320):106320.

9. Verma S, Bhatta M, Davies M, et al. Effects of once-weekly semaglutide 2.4 mg on C-reactive protein in adults with overweight or obesity (STEP 1, 2, and 3): Exploratory analyses of three randomised, double-blind, placebo-controlled, phase 3 trials. EClinicalMedicine. 2023;55(101737):101737.

10. Jensen MD, Ryan DH, Apovian CM, et al. 2013 AHA/ACC/TOS guideline for the management of overweight and obesity in adults: a report of the American College of Cardiology/American Heart Association Task Force on Practice Guidelines and The Obesity Society. J Am Coll Cardiol. 2014;63(25 Pt B):2985-3023.

11. Zhang Z, Li X, Wu X, Qiu H, Shi H, written on behalf of AME Big-Data Clinical Trial Collaborative Group. Propensity score analysis for time-dependent exposure. Ann Transl Med. 2020;8(5):246.

12. Benedetto U, Head SJ, Angelini GD, Blackstone EH. Statistical primer: propensity score matching and its alternatives. Eur J Cardiothorac Surg. 2018;53(6):1112–1117.

13. Shao S, Zhang X, Xu Q, Pan R, Chen Y. Emerging roles of Glucagon like peptide-1 in the management of autoimmune diseases and diabetes-associated comorbidities. Pharmacol Ther. 2022;239(108270):108270.

14. Wang Q, Anthony DD. Glucagon-like peptide-1 receptor analog use is associated with reduced thromboembolic events compared with dipeptidyl peptidase-4 inhibitors in rheumatoid arthritis patients: A global retrospective cohort study. Clin Rheumatol. Published online September 27, 2025. doi:10.1007/s10067-025-07709-0

15. Galli M, Benenati S, Laudani C, et al. Cardiovascular effects and tolerability of GLP-1 receptor agonists: A Systematic Review and meta-Analysis of 99,599 patients. J Am Coll Cardiol. Published online August 27, 2025. doi:10.1016/j.jacc.2025.08.027

16. Ussher JR, Drucker DJ. Glucagon-like peptide 1 receptor agonists: cardiovascular benefits and mechanisms of action. Nat Rev Cardiol. 2023;20(7):463–474.

17. Zhang S, Sidra F, Alvarez CA, Kinaan M, Lingvay I, Mansi IA. Healthcare utilization, mortality, and cardiovascular events following GLP1-RA initiation in chronic kidney disease. Nat Commun. 2024;15(1):10623.

18. Scott KW, Liu A, Chen C, et al. Healthcare spending in U.S. emergency departments by health condition, 2006-2016. PLoS One. 2021;16(10):e0258182.

19. Karacabeyli D, Lacaille D, Lu N, et al. Mortality and major adverse cardiovascular events after glucagon-like peptide-1 receptor agonist initiation in patients with immune-mediated inflammatory diseases and type 2 diabetes: A population-based study. PLoS One. 2024;19(8):e0308533.

20. Yen FS, Hou MC, Wei JCC, Shih YH, Hwu CM, Hsu CC. Effects of glucagon-like peptide-1 receptor agonists on liver-related and cardiovascular mortality in patients with type 2 diabetes. BMC Med. 2024;22(1):8.

21. Baviera M, Genovese S, Lepore V, et al. Lower risk of death and cardiovascular events in patients with diabetes initiating glucagon-like peptide-1 receptor agonists or sodium-glucose cotransporter-2 inhibitors: A real-world study in two Italian cohorts. Diabetes Obes Metab. 2021;23(7):1484–1495.

